# Lymphopenia predicts disease severity of COVID-19: a descriptive and predictive study

**DOI:** 10.1101/2020.03.01.20029074

**Authors:** Li Tan, Qi Wang, Duanyang Zhang, Jinya Ding, Qianchuan Huang, Yi-Quan Tang, Qiongshu Wang, Hongming Miao

**Author notes:** These authors contributed equally to this work (Li Tan, Qi Wang). Correspondence to: **Hongming Miao**, No. 30 Gaotanyan Street, Shapingba, Chongqing 400038, People’s Republic of China; **Qiongshu Wang**, No. 627 Wuluo Road, Wuchang District, Wuhan, Hubei province, 430015, People’s Republic of China.

## Abstract

**Background:** Coronavirus disease-2019 (COVID-19) is a rapidly escalating epidemic caused by SARS-CoV-2. Identification of a simple and effective indicator to assess disease severity and prognosis is urgently needed.

**Methods:** Dynamic changes of blood lymphocyte percentage (LYM%) in 15 death cases, 15 severe cases as well as 40 moderate cases of COVID-19 patients were retrospectively analyzed. A Time-LYM% model (TLM) was established according to the descriptive studies and was validated in 92 hospitalized cases.

**Results:** Results from death and severe cases showed that LYM% in blood tests were inversely associated with the severity and prognosis of COVID-19. LYM% in moderate type of patients with COVID-19 remained higher than 20% 10-12 days after symptom onset. In contrast, LYM% was lower than 20% in severe cases. However, LYM% in severe cases was higher than 5% 17-19 days after the onset of the disease, while it fell below 5% in death cases. Accordingly, we established a Time-LYM% model (TLM), which was validated as an independent criterion of disease classification in another 92 hospitalized patients with COVID-19.

**Conclusion:** Lymphopenia can be used as an indicator of disease severity and prognosis of COVID-19 patients. TLM is worth of application in the clinical practice.

## Introduction

Coronaviruses are a large family of viruses that cause both common cold and serious respiratory illnesses, such as Middle East Respiratory Syndrome (MERS) and Severe Acute Respiratory Syndrome (SARS)^1-3^. An outbreak of an unknown infectious pneumonia has recently occurred in Wuhan, China^4^. The pathogen of the disease was quickly identified as a novel coronavirus (SARS-CoV-2, severe acute respiratory syndrome coronavirus 2), and the disease was named coronavirus infection disease-19 (COVID-19)^5^. the virus has so far caused 78959 confirmed cases and 2791 deaths in China according to WHO. COVID-19 has been spreading in many countries such as Japan, Korea, Singapore, Iran and Italia The clinical manifestation of COVID-19 include fever, cough, fatigue, muscle pain, diarrhea, and pneumonia, which can developed to acute respiratory distress syndrome, metabolic acidosis, septic shock, coagulation dysfunction, and organ failure such as liver, kidney and heart failure^4,6,7^. Unfortunately, There is no effective medication other than comprehensive support. However, the mild type of COVID-19 patients can recover shortly after appropriate clinical intervention. The moderate type patients, especially the elderly or the ones with comorbidity, can worsen and became severe, indicating high mortality^6,7^. However, efficient indicators for the disease severity, therapeutic response and disease outcome are still incompletely investigated. Once such indicators are present, reasonable medication and care can be inclined, which is believed to significantly reduce the mortality of severe patients.

Routine examinations include complete blood count, coagulation profile, and serum biochemical test (including renal and liver function, creatine kinase, lactate dehydrogenase, and electrolytes). Complete blood count is the most available, efficient and economic examination. This study aimed to retrospect and analyze the time-courses of complete blood count of cured and dead patients, in order to obtain key indicators of disease progression and outcome and to provide guidance for subsequent clinical practice.

## Methods

### Patients

In this study, all cases were taken from the General Hospital of Central Theater Command (Wuhan, Hubei province, People’s Republic of China), one of designated hospital for the COVID-19 by local authority. This study was approved by the Ethics Committee of the hospital. All subjects signed informed consent forms at admission to hospital. In this study, 162 patients including the hospitalized, discharged and died in our hospital between January 14, 2020 and February 25, 2020 were investigated. Among them, 95 cases were moderate, 40 cases of which were cured and discharged from the hospital. There were 39 cases of severe type and 28 cases of critically ill type. All patients were confirmed by viral detections using quantitative RT-PCR, which ruled out infection by other respiratory viruses such as influenza virus A, influenza virus B, coxsackie virus, respiratory syncytial virus, parainfluenza virus and enterovirus by the same time. All cases were diagnosed and classified according to the New Coronavirus Pneumonia Diagnosis Program (5th edition) published by the National Health Commission of China^8^. Clinical manifestations consist of four categories, mild, moderate, severe and critical. The mild clinical symptom were mild with no pulmonary inflammation on imaging. The moderate is the overwhelming majority, showing symptoms of respiratory infections such as fever, cough, and sputum, and pulmonary inflammation on imaging; when symptoms of dyspnea appear, including any of the following: shortness of breath, RR ≥ 30bpm, blood oxygen saturation ≤ 93% (at rest), PaO_2_ / FiO_2_ ≤ 300 mmHg, or pulmonary inflammation that progresses significantly within 24 to 48 hours> 50%, it was classified as severe; respiratory failure, shock, and organ failures that require intensive care were critically ill. Among them, mild patients were not admitted in this designated hospital.

### Data collection

In this study, the basic information, complete blood count, coagulation profile, and serum biochemical test (including renal and liver function, creatine kinase, lactate dehydrogenase, and electrolytes) and disease outcome of all included patients were collected.

### Statistical methods

In this study, GraphPad 6.0 software was used for data statistics and mapping. The presentation of dynamic changes of blood markers is descriptive. The consistence between Guideline and TLM-based disease classification was tested using kappa statistic. Kappa≧0.75 indicates a high consistence and 0.75>Kappa≧0.4 indicates a general consistence.

## Results

### Changes in blood test results from COVID-19 patients

In order to explore the relationship between tested blood markers and disease conditions in COVID-19 patients, we first randomly selected 5 death cases and monitored dynamic changes in blood tests for each patient from disease onset to death. Although course of disease in each patient was different, inter-day variations of most parameters studied are fairly constant among all five patients. Common blood tests showed that fluctuations of red blood cells and neutrophils were not obvious (Fig. 1A, D). The percentage of white blood cells is increased during disease progression in four out of five patients (Fig. 1B). Platelets count, monocyte percentage, and lymphocyte percentage (LYM%) all showed the downward trend (Fig.1C, E and F). Among all factors, LYM% showed the most significant and consistent trend (Fig.1F), indicating that this indicator might reflect the disease progression. In addition, we also investigated the indicators reflecting liver, kidney and myocardial functions. Significant changes in these indicators were usually accompanied by organ failure and cytokine storm syndromes (data not shown), which occurred at the end of disease course and thus could not reflect disease progression. Therefore, we only focused on lymphopenia in COVID-19 in the present study.

**Figure 1.**
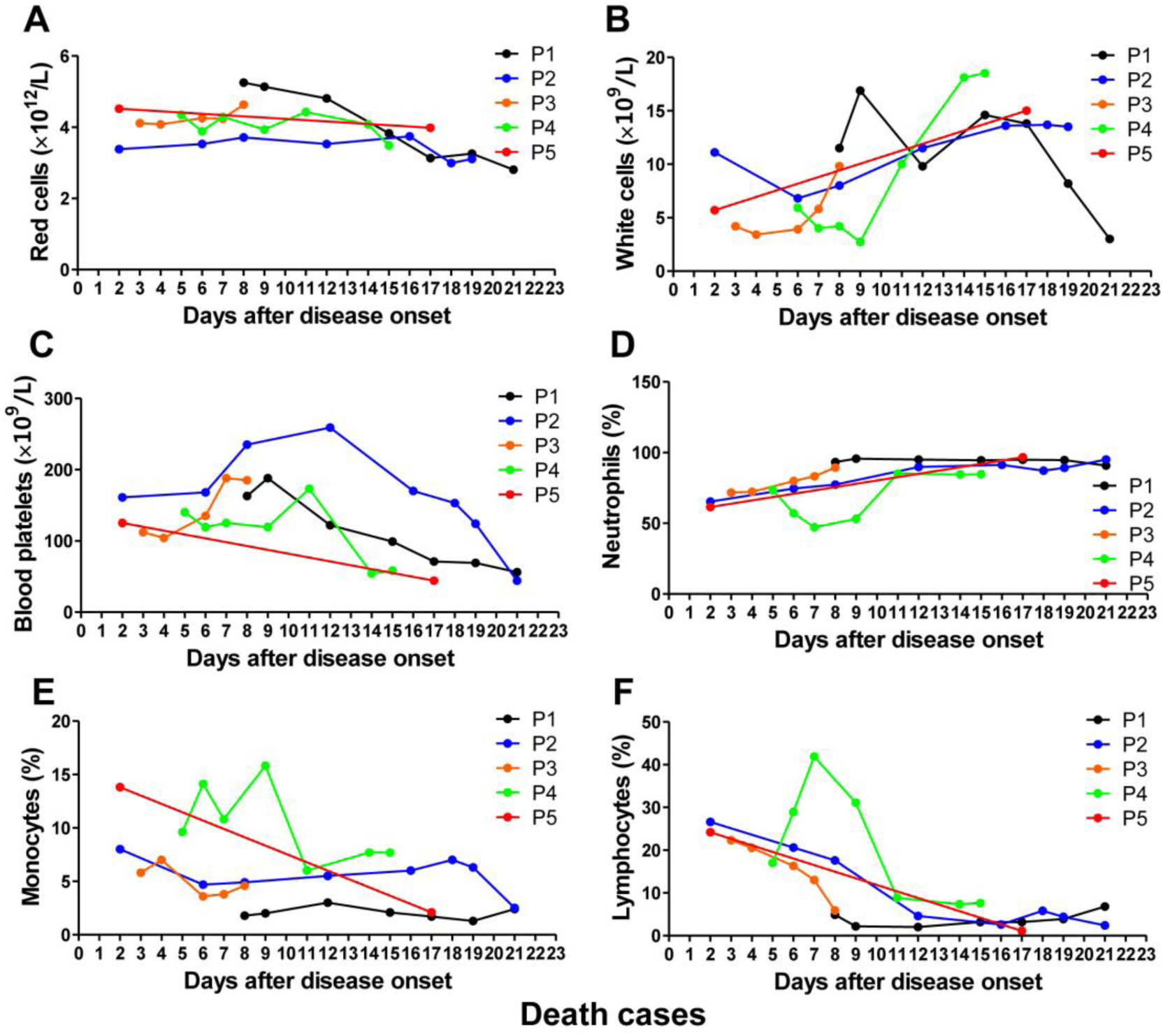
Dynamic changes of routine indicators in blood tests of COVID-19 patients. **(A-F)** Dynamic changes of red cells (**A**), white cells (**B**), platelets (**C**), neutrophils (**D**), monocytes (**E**) and lymphocytes (**F**) in the routine blood tests of COVID-19-caused death cases. The descriptive curve of individual patient P1, P2, P3, P4 and P5 was displayed. (n=5)

### Low LYM% is a predictor of prognosis in COVID-19 patients

To further confirm the relationship between blood LYM% and patient’s condition, we increased our sample size to 12 death cases (mean age: 76 years; average therapeutic time: 20 days) (Supplementary Table 1). Most cases showed that LYM% was reduced to lower than 5% within 2 weeks after disease onset (Fig. 2A). We also randomly selected 7 cases (mean age: 35 years, average therapeutic time: 35 days) with severe symptoms and cured outcome (Supplementary Table 2) and 11 cases (mean age: 49; average therapeutic time: 26 days) with moderate symptoms and cured outcomes (Supplementary Table 3). LYM% of severe patients fell down initially and then rose to higher than 10% until discharged (Fig. 2B). In contrast, LYM% of moderate patients fluctuated very little after disease onset and was higher than 20% when discharged (Fig. 2C). These results suggest that lymphopenia is a predictor of prognosis in COVID-19 patients.

**Figure 2.**
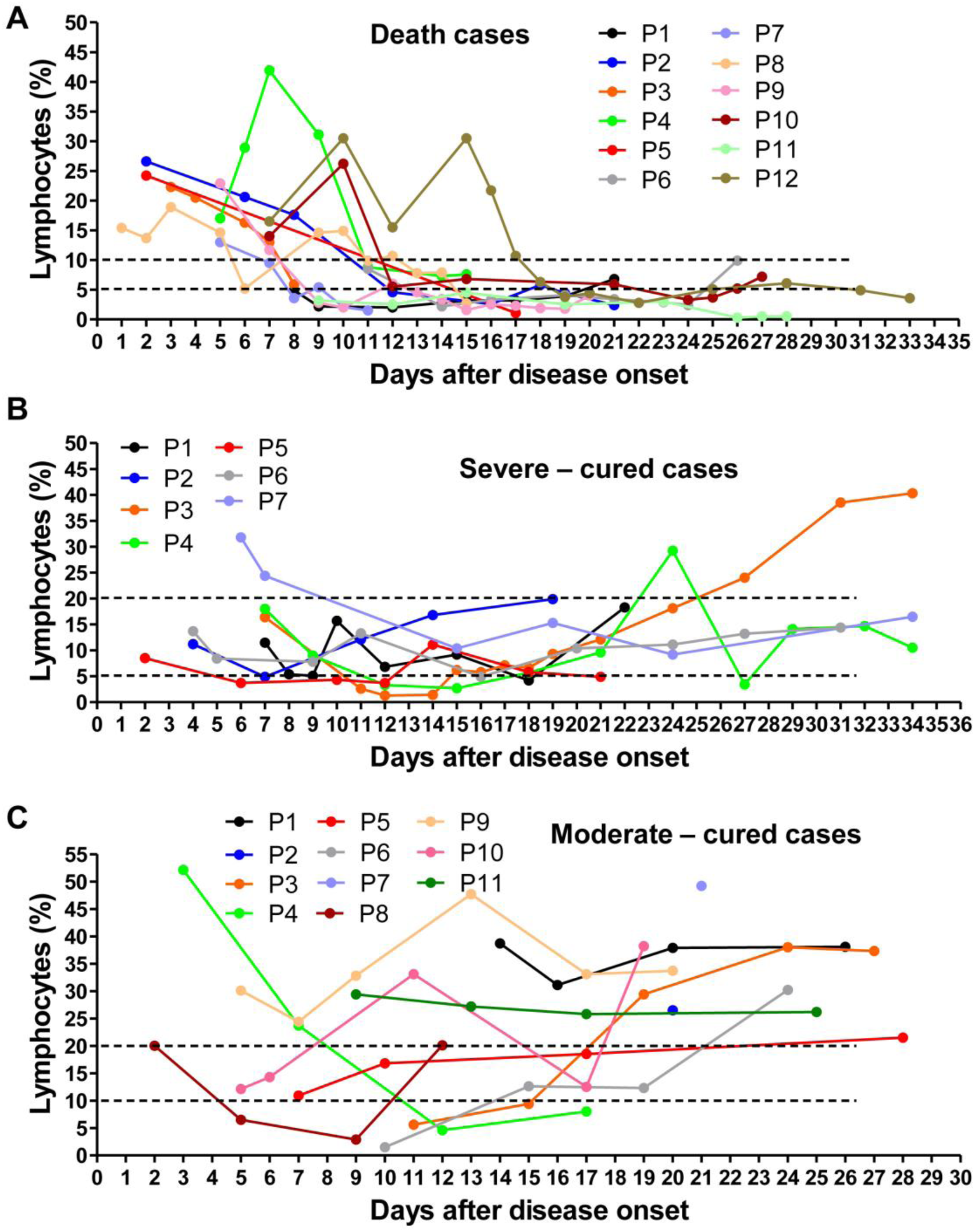
Dynamic changes of LYM% in the death, severe-cured and moderate-cured patients with COVID-19. **(A)** Dynamic changes of LYM% in the death cases (n=12). **(B)** Dynamic changes of LYM% in the severe-cured cases (n=7). **(C)** Dynamic changes of LYM% in the moderate-cured cases (n=11). The data of each patient was showed by a descriptive curve. Severe-cured: severe type with a cured outcome; moderate-cured: moderate type with a cured outcome

### Establishment of a Time-LYM% model

By summarizing all the death cases, severe-cured cases and moderate-cured cases in our hospital to depict the time-LYM% curve (Fig. 3A), we established a Time-LYM% model (TLM) for disease classification and predicting the disease outcome of COVID-19 patients (Fig. 3B) and. We defined TLM as follows: patients have varying LYM% after the onset of COVID-19. At the 1^st^ time point (TLM-1) of 10-12 days after symptom onset, patients with LYM% > 20% are classified as moderate type, who can recover quickly. Patients with LYM% < 20% are initially classified as severe type. At the 2^nd^ time point (TLM-2) of 17-19 days after symptom onset, patients with LYM% > 20% are in recovery; patients with 5% < LYM% < 20% are still in danger and in need of supervision; patients with LYM% < 5% become critically ill with high mortality and need intensive care.

**Figure 3.**
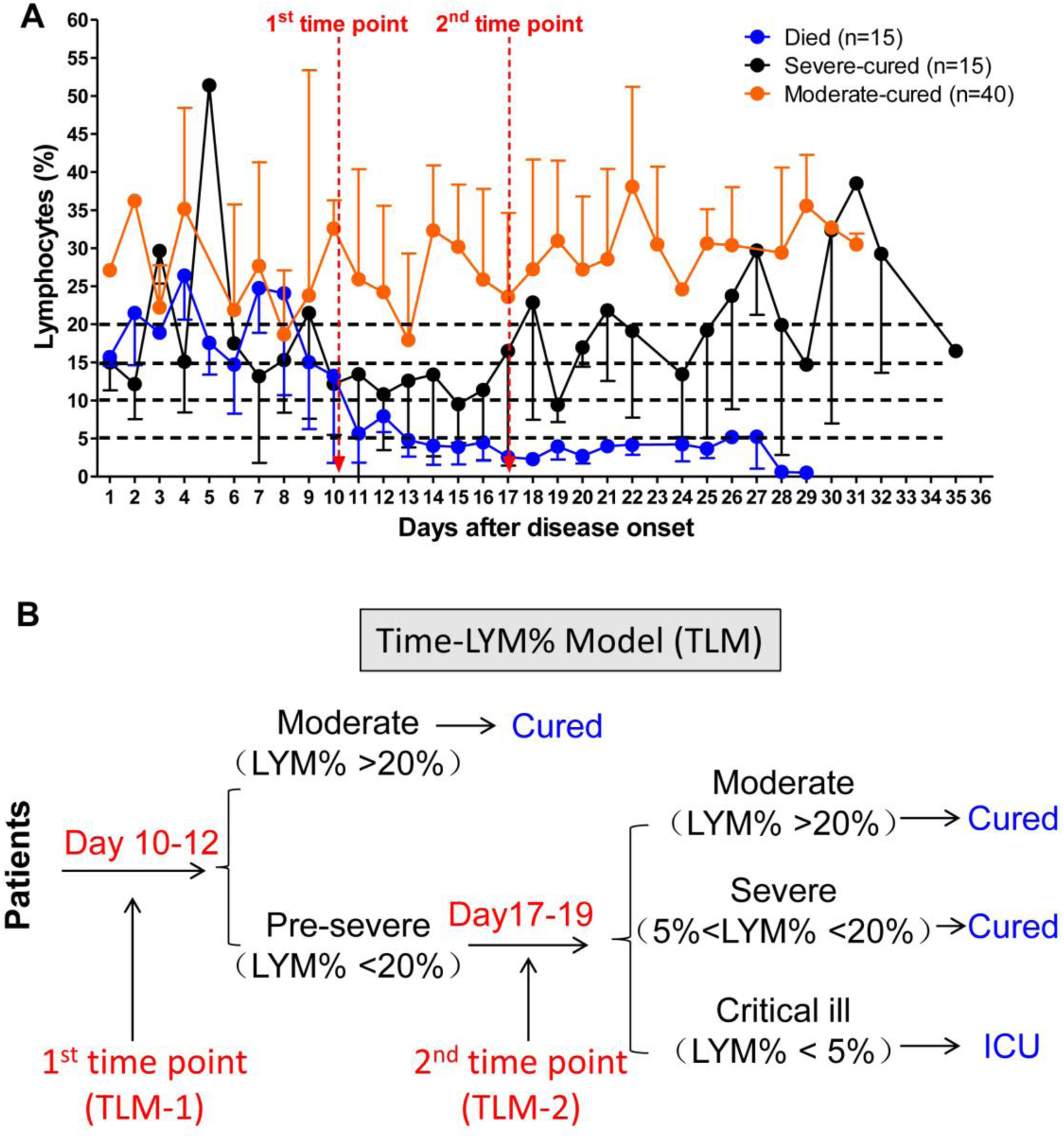
Establishment of Time-LYM% model (TLM) **(A)** Dynamic changes of LYM% in the death cases (n=15), severe-cured cases (n=15) and moderate-cured cases (n=40). Data were showed as means ± s.e.ms. Two cut-off time points of these three curves were set as 1^st^ time point (day 10-12) and 2^nd^ time point (day 17-19). **(B)** Description of TLM: 1^st^ time point (TLM-1) and 2^nd^ time point (TLM-2) are defined as day 10-12 and day 17-19 from symptom onset, respectively. The confirmed COVID-19 patients with LYM%>20% at TLM-1 are classified as moderate type and the ones with LYM%<20% at TLM-1 are suggested as pre-severe type which need to be further distinguished at TLM-2. If LYM%>20% at TLM-2, those pre-severe patients are reclassified as moderate. If 5%<LYM%<20% at TLM-2, the pre-severe patients are indeed typed as severe. If LYM%<5% at TLM-2, those patients are suggested as critically ill. The moderate and severe types are curable, while the critically ill type needing intensive care has a poor prognosis.

### Application and validation of TLM

We further collected information from all COVID-19 patients who were admitted to hospital, and validated the consistency between TLM and the existing guideline. As shown in Figure 4, 92 COVID-19 patients were currently hospitalized in light of the classification criteria of the Chinese New Coronavirus Pneumonia Diagnosis Program (5th edition): 55 moderate patients, 24 severe patients and 13 critically ill patients. According to the TLM we proposed, LYM% in 24 out of 55 moderate cases was lower than 20% at TLM-1; LYM% of all these patients was above 5%, indicating that these patients would recover soon. Regarding other 24 patients with severe symptoms, LYM% at the TLM-1 was lower than 20% in 20 out of 24 cases. LYM% at TLM-2 in 6 cases was less than 5%, indicating a poor prognosis. LYM% in 12 out of 13 critically ill patients at the TLM-1 was lower than 20%. LYM% of these patients at TLM-2 in 6 cases was lower than 5%, suggesting a poor prognosis (Fig. 4A). Furthermore, with kappa statistic test, we further verified the consistence between TLM and the existing guideline in disease typing (Fig. 4B).

**Figure 4.**
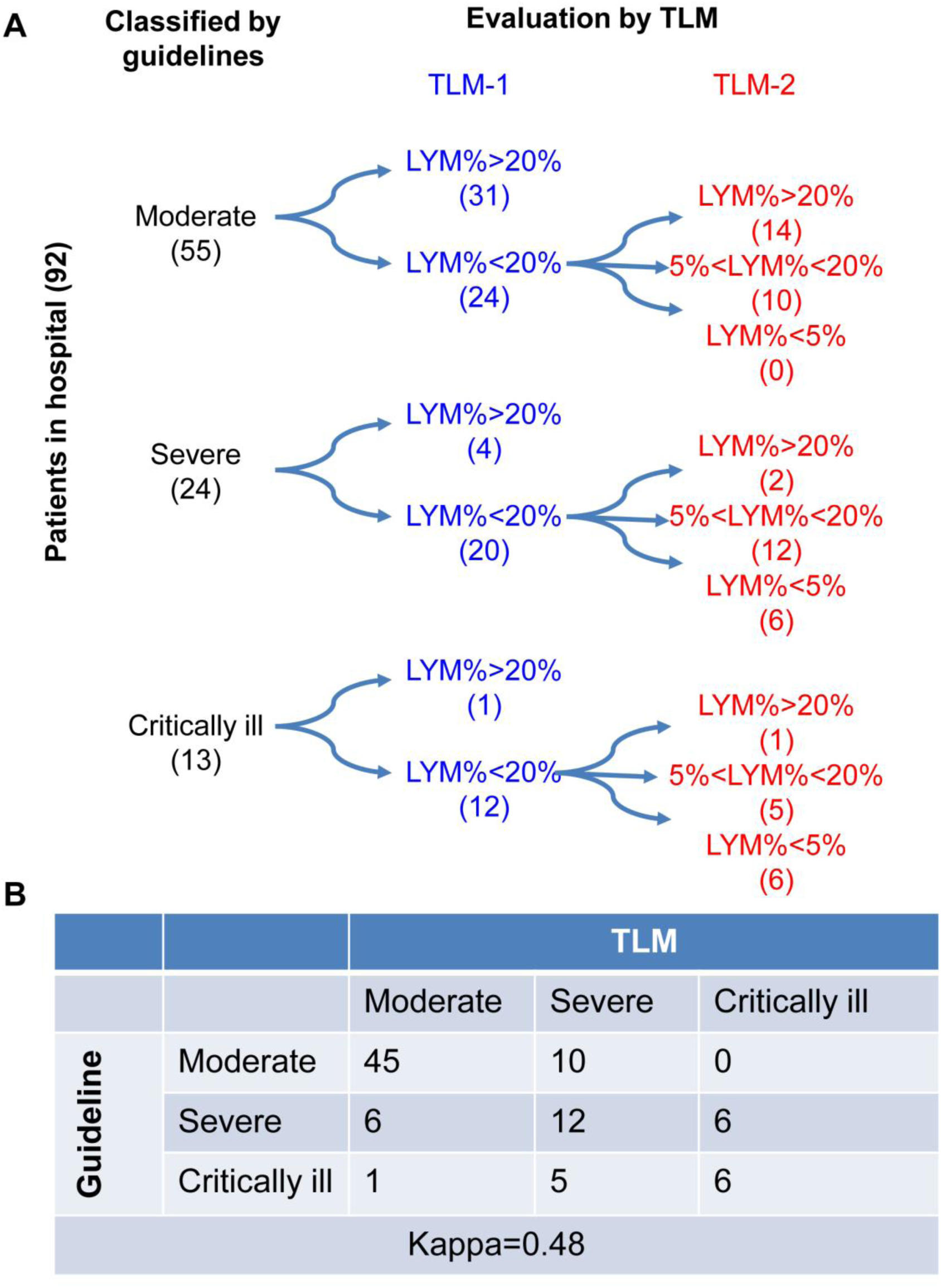
Validation of the reliability of TLM in hospitalized patients with COVID-19. **(A)** 92 COVID-19 patients were currently hospitalized in light of the classification criteria of the New Coronavirus Pneumonia Diagnosis Program (5th edition) (5th edition): 55 patients with moderate type, 24 patients with severe type and 13 patients with critically ill type. At TLM-1, LYM% in 24 out of 55 moderate cases was lower than 20%; At TLM-2, LYM% in all 24 patients was above 5%, indicating that these patients would be curable. Regarding other 24 patients with severe symptoms, LYM% at TLM-1 was lower than 20% in 20 out of 24 cases. LYM% at TLM-2 in 6 cases was less than 5%, indicating a poor prognosis. In 12 out of 13 critically ill patients, LYM% at TLM-1 was lower than 20%. LYM% at TLM-2 in 6 cases was lower than 5%, suggesting a poor prognosis. **(B)** The consistence between Guideline and TLM-based disease classification in (**A**) was tested using kappa statistic. Kappa = 0.48

## Discussion

By retrospectively tracking the dynamic changes of LYM% in death cases and cured cases, this study suggests that lymphocyte count is an effective and reliable indicator for disease classification and prognosis in COVID-19 patients. We also established TLM innovatively and verified its reliability in a considerable number of hospitalized COVID-19 patients. A high correlation of blood lymphocytes with the disease progression suggested that lymphocyte deficiency or incapacity is the key cellular pathology of COVID-19. The protection, maintenance or promotion of lymphocyte levels might have a good effect on the prevention and treatment of COVID-19.

### LYM% indicates disease severity of COVID-19

The classification of disease severity in COVID-19 is very important for the grading treatment of patients. In particular, when the outbreak of an epidemic occurs and medical resources are relatively scarce, it is necessary to conduct grading severity and treatment, thus optimizing the allocation of rescue resources, and prevent the occurrence of overtreatment or undertreatment. According to the latest 5^th^ edition of the national treatment guideline, COVID-19 can be classified into four types. Pulmonary imaging is the main basis of classification, and other auxiliary examinations are used to distinguish the severity. Blood tests are easy, fast and cost-effective. However, none of the indicators in blood tests were included in the classification criteria. This study suggested that LYM% can be used as a reliable indicator to classify the moderate, severe and critical ill types independent of any other auxiliary indicators.

### LYM% indicates the outcome of COVID-19 patients

During the development of mild disease into severe condition, the proportion of lymphocytes in the blood gradually decreased and maintained at a low level. By the time the disease began to improve, LYM% in the blood gradually rose to normal or nearly normal levels. Patients with persistently low levels of blood lymphocytes, especially less than 5%, often had a poor prognosis. Therefore, we suggested that LYM% should be used as an indicator for evaluating the effectiveness of clinical drugs or therapies. We suggest more medical staff apply and improve our proposed TLM to give patients more timely and appropriate treatments.

### Analysis of possible reasons for lymphopenia in COVID-19

Lymphocytes play a decisive role in maintaining immune homeostasis and inflammatory response throughout the body. Understanding the mechanism of reduced blood lymphocyte levels is expected to provide an effective strategy for the treatment of COVID-19. We speculated four potential mechanisms leading to lymphocyte deficiency. (1) The virus might directly infect lymphocytes, resulting in lymphocyte death. Lymphocytes express the coronavirus receptor ACE2 and may be a direct target of viruses.^9^ (2) The virus might directly destroy lymphatic organs. Acute lymphocyte decline might be related to lymphocytic dysfunction, and the direct damage of novel coronavirus virus to organs such as thymus and spleen cannot be ruled out. This hypothesis needs to be confirmed by pathological dissection in the future. (3) Inflammatory cytokines continued to be disordered, perhaps leading to lymphocyte apoptosis. Basic researches confirmed that TNFα, IL-6 and other pro-inflammatory cytokines could induce lymphocyte deficiency^10^. (4) Inhibition of lymphocytes by metabolic molecules produced by metabolic disorders, such as hyperlactic acidemia. The severe type of COVID-19 patients had elevated blood lactic acid levels, which might suppress the proliferation of lymphocytes^11^. Multiple mechanisms mentioned above or beyond might work together to cause lymphopenia, and further research is needed.

### Shortcomings

The clinical data in the present study came from a single center and the sample size was limited. The TLM we established was based on the specific treatment condition in the hospital and might not be fully applicable to some patients who were treated in light of a different guideline.

## Conclusion

Lymphopenia is an effective and reliable indicator of severity and hospitalization in COVID-19 patients. We suggest that the TLM should be included in the diagnosis and therapeutic guidelines of COVID-19.

## Data Availability

All the data that support the findings of this study are available within the article and its Supplementary file or from the corresponding author upon reasonable request. The supplementary file includes 3 Tables.

## Acknowledgements

We thank all the doctors, nurses, public health workers, and researchers for the braveness in fighting against SARS-CoV-2 and the efforts to save the life of COVID-19 patients. This work was supported by the award numbers cstc2017jcyjBX0071 (H.M.) from the Foundation and Frontier Research Project of Chongqing and T04010019 (H.M.) from Chongqing youth top talent project.

## Contributions

Li Tan, Qi Wang, Qiongshu Wang were responsible for the collection and summary of clinical cases. Duanyang Zhang, Jinya Ding, Qianchuan Huang and Yi-Quan Tang participated in discussion. Yi-Quan Tang and Qi Wang contributed to language polishing. Hongming Miao was responsible for conceptual design, data analysis, manuscript writing and submission.

## References

1. Kuiken T, Fouchier RA, Schutten M, et al. Newly discovered coronavirus as the primary cause of severe acute respiratory syndrome. Lancet 2003; 362(9380): 263–70.

2. Drosten C, Gunther S, Preiser W, et al. Identification of a novel coronavirus in patients with severe acute respiratory syndrome. N Engl J Med 2003; 348(20): 1967–76.

3. de Groot RJ, Baker SC, Baric RS, et al. Middle East respiratory syndrome coronavirus (MERS-CoV): announcement of the Coronavirus Study Group. J Virol 2013; 87(14): 7790–2.

4. Chen N, Zhou M, Dong X, et al. Epidemiological and clinical characteristics of 99 cases of 2019 novel coronavirus pneumonia in Wuhan, China: a descriptive study. Lancet 2020; 395(10223): 507–13.

5. Zhu N, Zhang D, Wang W, et al. A Novel Coronavirus from Patients with Pneumonia in China, 2019. N Engl J Med 2020; 382(8): 727–33.

6. Wang D, Hu B, Hu C, et al. Clinical Characteristics of 138 Hospitalized Patients With 2019 Novel Coronavirus-Infected Pneumonia in Wuhan, China. JAMA 2020.

7. Huang C, Wang Y, Li X, et al. Clinical features of patients infected with 2019 novel coronavirus in Wuhan, China. Lancet 2020; 395(10223): 497–506.

8. Zu ZY, Jiang MD, Xu PP, et al. Coronavirus Disease 2019 (COVID-19): A Perspective from China. Radiology 2020: 200490.

9. Xu H, Zhong L, Deng J, et al. High expression of ACE2 receptor of 2019-nCoV on the epithelial cells of oral mucosa. Int J Oral Sci 2020; 12(1): 8.

10. Liao YC, Liang WG, Chen FW, Hsu JH, Yang JJ, Chang MS. IL-19 induces production of IL-6 and TNF-alpha and results in cell apoptosis through TNF-alpha. J Immunol 2002; 169(8): 4288–97.

11. Fischer K, Hoffmann P, Voelkl S, et al. Inhibitory effect of tumor cell-derived lactic acid on human T cells. Blood 2007; 109(9): 3812–9.

